# Patterns of Physical Activity Over Time in Older Patients Rehabilitating after Hip Fracture Surgery: A Preliminary Study

**DOI:** 10.1101/2022.06.09.22276191

**Authors:** Dieuwke van Dartel, Ying Wang, Johannes H. Hegeman, Marloes Vermeer, Miriam M.R. Vollenbroek-Hutten

## Abstract

**Purpose:** To investigate patterns of continuously monitored physical activity in older patients rehabilitating after hip fracture surgery and the association with patient characteristics.

**Methods:** Physical activity of surgically treated hip fracture patients (≥70 years) was continuously measured during rehabilitation at a skilled nursing home using an accelerometer. The intensity of physical activity per day was calculated to describe patients’ activity. Physical activity patterns of overall activity, overall variability, and day-to-day variability were investigated. Two experts in geriatric rehabilitation visually identified unique physical activity patterns for each aspect. Eighteen healthcare professionals independently classified each patient in one of the predefined patterns for each aspect. Differences between physical activity patterns and patient characteristics were assessed using a Kruskal-Wallis or Fisher’s Exact Test.

**Results:** Data from 66 patients were used. Six unique patterns were identified for overall activity and overall variability, and five for day-to-day variability. The most common pattern of overall activity and day-to-day variability first slowly increased, then steeply increased, and subsequently flattened (n=23,34.8%). For overall variability, the most common pattern first slowly increased, then steeply increased, then decreased and lastly increased (n=14,21.2%). Differences in functionality at rehabilitation admission, measured with the Barthel Index, and duration of rehabilitation stay were found between patterns of physical activity.

**Conclusions:** This preliminary study showed different patterns of physical activity among older hip fracture patients during rehabilitation. Functionality at rehabilitation admission and duration of rehabilitation stay were associated with these different patterns. Differences in physical activity patterns emphasize the importance of personalized hip fracture treatment

## INTRODUCTION

Hip fractures have a major impact on the physical functioning of older patients, i.e. the mobility and the ability to perform Activities of Daily Living (ADL). A high number of older patients do not recover to their premorbid level of functioning after a hip fracture, and full functional recovery can take up to 12 months.^1–3^ Being physically active during rehabilitation after hip fracture surgery is highly relevant in the recovery process of older patients; more physical activity is associated with higher functional outcomes and a decreased risk of future falls.^4–11^ However, restoring physical activity during rehabilitation after hip fracture surgery differs among patients and could be influenced by factors like fear of falling, pain, age, gender, or comorbidities.^7–14^ Currently, still little is known about how older patients evolve in their physical activity over time during hip fracture rehabilitation and about the patterns of physical activity over time. More insight about the evolution and patterns of physical activity over time, and the factors associated with these patterns, is therefore considered highly important. With these insights, healthcare processes can be improved, clinical staff can adopt a more pro-active treatment policy and it can stimulate patient motivation.

Commonly used methods for physical activity monitoring during rehabilitation such as Patient Reported Outcome Measures (PROMs), clinimetric tests, or lab-based gait analysis, are not suitable to gain insight in an older patient’s daily level of physical activity.^15–17^ Shortcomings such as subjective and static measurements, expensive equipment, and specialized laboratory set-ups are reasons for this low suitability.^15–18^ Therefore, an objective and continuous way of monitoring physical activity over time during rehabilitation is needed. Wearable devices that can perform this way of physical activity monitoring with minimal inconvenience of the user are expected to be useful to monitor the patterns of physical activity in older patients after hip fracture surgery.^16, 17, 19–21^ Continuous physical activity information can be used to assess an older patients’ progress and assist healthcare professionals in providing personalized feedback.^21^

Currently, only one study was found assessing the trajectories of recovery in older patients after hip fracture surgery by continuously monitoring physical activity for four months using an activity monitor.^22^ They showed that the average physical activity was significantly higher in older patients who gained full mobility four months after hip fracture surgery compared with older patients who did not. However, in this study, only a limited number of older patients were included (n=13). Furthermore, the individual heterogeneity was not investigated for the specific patterns of physical activity of each older patient, so factors associated with the trajectories of recovery are still unknown. Therefore, this preliminary study aims a) to investigate the patterns of physical activity over time in older patients rehabilitating after hip fracture surgery by continuously monitoring physical activity using a wearable device, and b) to assess whether the patterns of physical activity over time were associated with patient characteristics.

## METHODS

### Study Design

This study was a preliminary study with a longitudinal observational design, which was conducted in the period from January 2019 until June 2021 as part of a larger project called the “Up&Go after a hip fracture” project. The Up&Go project focuses on the optimization of the rehabilitation of older patients after hip fracture surgery. The project was approved by the Medical Research Ethics Committee (MREC) Twente and by the institutional review board of Ziekenhuisgroep Twente (ZGT), The Netherlands. The MREC Twente considered the project as not subject to the Medical Research Involving Human Subjects Act (WMO). All patients gave written informed consent to participate.

### Study Population

Older patients were enrolled in this study when they were 70 years or older, surgically treated for a hip fracture at the department of Trauma Surgery of ZGT, and temporarily admitted for geriatric rehabilitation to one of the three collaborating skilled nursing homes (TriviumMeulenbeltZorg, Carintreggeland, and ZorgAccent). Exclusion criteria were severe cognitive impairment (i.e. diagnosed with dementia), a total hip replacement, a pathological or periprosthetic fracture, plaster allergy, terminal illness or contact isolation. Patients were enrolled in this study one day before discharge from the hospital to the geriatric rehabilitation department of one of the skilled nursing homes.

### Study Measurements

Physical activity of all enrolled patients was continuously monitored during their entire stay at the rehabilitation department at the skilled nursing home using a MOX sensor (Maastricht Instruments BV, The Netherlands). The MOX is a comfortable, small, single-unit, waterproof device which was attached to the upper thigh of the older adult, approximately 10 cm above the knee, using a biocompatible body attachment patch from Maastricht Instruments BV (Figure 1). The MOX contains a tri-axial accelerometer to continuously monitor physical activity and has a sample frequency of 25 Hz. Furthermore, it has a data storage of 1.5 GB and a battery life of seven days. Raw MOX acceleration data obtained during daytime (7.00 am to 10.00 pm) were used to analyze the intensity of physical activity. The intensity of physical activity was chosen as this metric can properly describe an older patient’s general physical activity level and is easy to interpret. Raw acceleration data were pre-processed using a moving average filter with a window size of 0.12 seconds and a fourth order Butterworth High Pass filter with a cut-off frequency of 1 Hz to remove noises and gravitational accelerations. The intensity of physical activity was calculated as the area under the curve of the accelerometer signals and expressed in activity counts. The intensity of physical activity was calculated per day for all older patients using Matlab (R2017b, MathWorks Inc., Natick, MA, USA). For more details about the data analysis see Appendix A.

**Figure 1:**
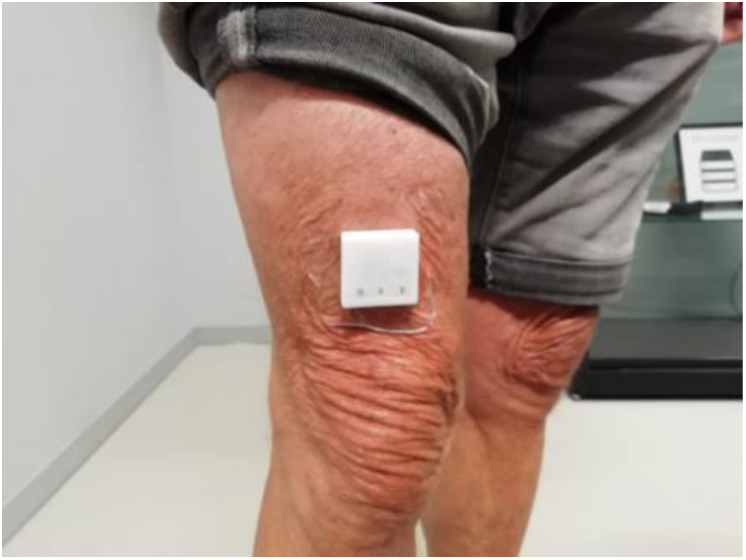
A MOX device attached to the upper thigh of a person, approximately 10 cm above the knee. The MOX was attached using a special plaster.

All enrolled patients received the usual rehabilitation care during their rehabilitation at the skilled nursing home consisting of multidisciplinary treatment by a nursing home physician, physiotherapist, occupational therapist, psychologist, dietician, and nursing staff. Furthermore, all older patients received physiotherapy sessions on all weekdays. Information was gathered about the following patient variables: age, gender, premorbid living situation, premorbid Katz index of independence in Activities of Daily Living (Katz-ADL), Pre-Fracture Mobility Score (PFMS), comorbidities (Charlson Comorbidity Index [CCI]), type of surgery, cognition (Montreal Cognitive Assessment [MoCA]), the level of functioning at admission to the rehabilitation department (Fracture Mobility Score [FMS], Functional Ambulation Categories [FAC], Barthel Index [BI], and weight bearing policy), complications during rehabilitation, and the level of functioning at discharge from the rehabilitation department (FMS, FAC, and BI). The Katz-ADL measured the level of independence in ADL and ranged from 0 (completely independent) to 6 (completely dependent).^23, 24^ The PFMS and FMS scored the level of mobility and ranged from 0 (fully mobile without aids) to 5 (no functional mobility).^25^ The CCI assessed the prognostic burden of comorbid diseases by assigning different weights to specific diseases.^26^ The MoCA assessed whether an older patient had a mild cognitive impairment or not.^27^ The MoCA ranged from 0 to 30 and a score of ≥ 26 was considered normal. The FAC scored the walking ability and ranged from 0 (not able to walk) to 5 (independent walking).^28^ The BI measured the level of independence in ADL and ranged from 0 (completely dependent) to 20 (completely independent).^29^

### Physical Activity Patterns

The patterns of three different aspects (A-C) of physical activity over time were investigated to gain insights into the patterns of overall physical activity over time and the patterns of variability in physical activity over time.

A. The pattern of overall intensity of physical activity over time, further referred as pattern of overall physical activity. For all patients, the overall intensity of physical activity per day was plotted against time to assess how patients evolve in physical activity during their rehabilitation (Figure 2A).
B. The pattern of variability in the overall intensity of physical activity between days over time, further referred as pattern of variability in overall physical activity. This pattern gives an indication of the variance in physical activity between days during rehabilitation. For this aspect, we calculated the variability in the overall intensity of physical activity with an overlapping, centered sliding window of 5 days. This variability was then plotted against each window to assess for patterns of variability in physical activity for all patients (Figure 2B).
C. The pattern of variability in the intensity of physical activity within each day over time, further referred as pattern of day-to-day variability. This pattern gives an indication of the variance in physical activity within each day during rehabilitation. The variability within each day was plotted against time to assess for patterns of the day-to-day variability (Figure 2C).

**Figure 2:**
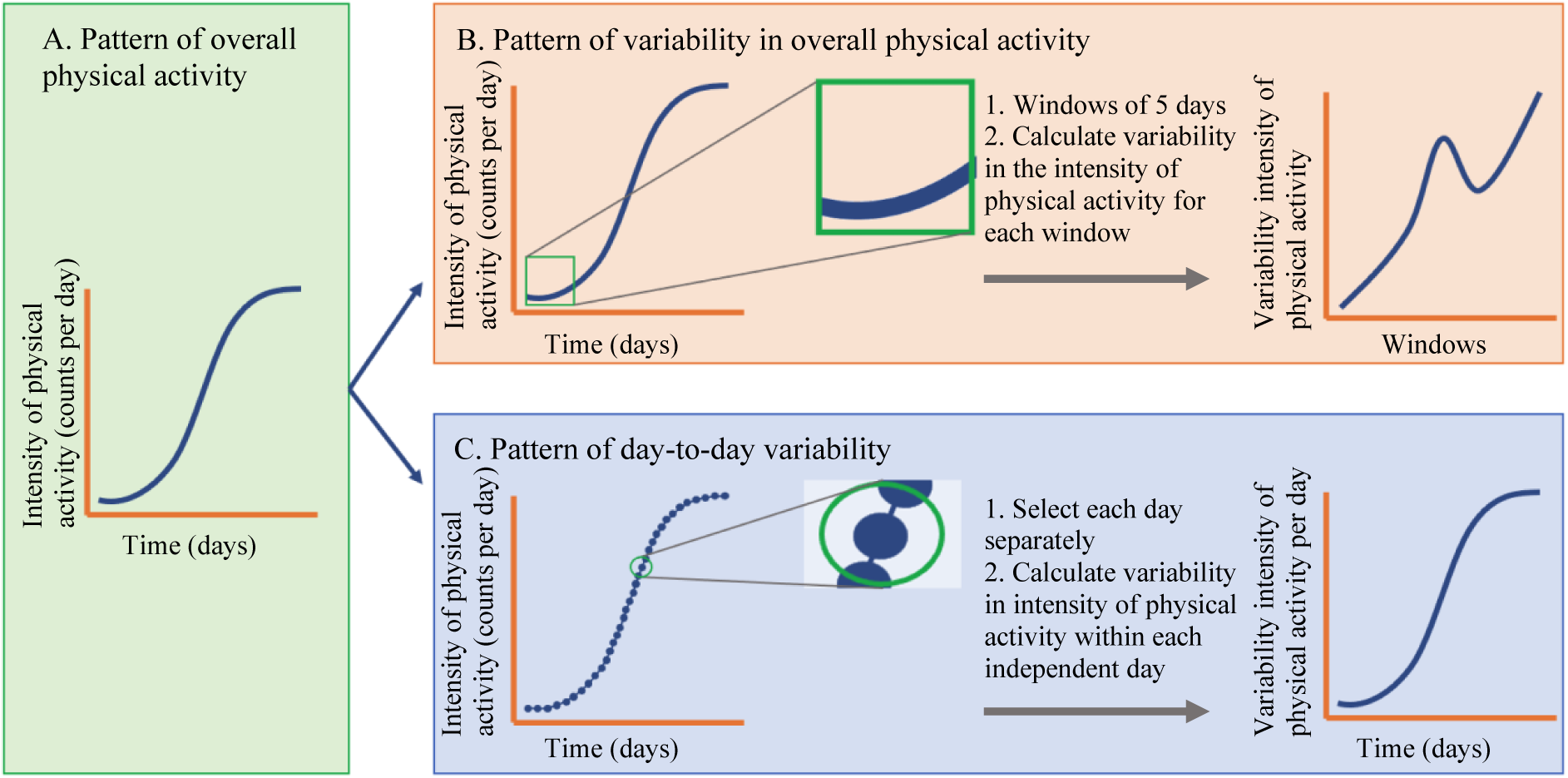
three different aspects of physical activity were assessed in this study. A. describes the pattern of overall physical activity, B. describes the pattern of variability in overall physical activity, and C. described the pattern of day-to-day variability.

All figures were smoothed with a Gaussian-weighted moving average smoothing filter to get the general trend of the physical activity patterns. Patterns of physical activity for each enrolled patient and each physical activity aspect (A-C) were then assessed based on visual analysis.

#### Identification of unique physical activity patterns

For all three physical activity aspects, two experts in the geriatric rehabilitation field, both nursing home physicians, identified unique patterns based on visual analysis and their experience in clinical practice. The unique patterns are shown in Figure 3A-C.

**Figure 3:**
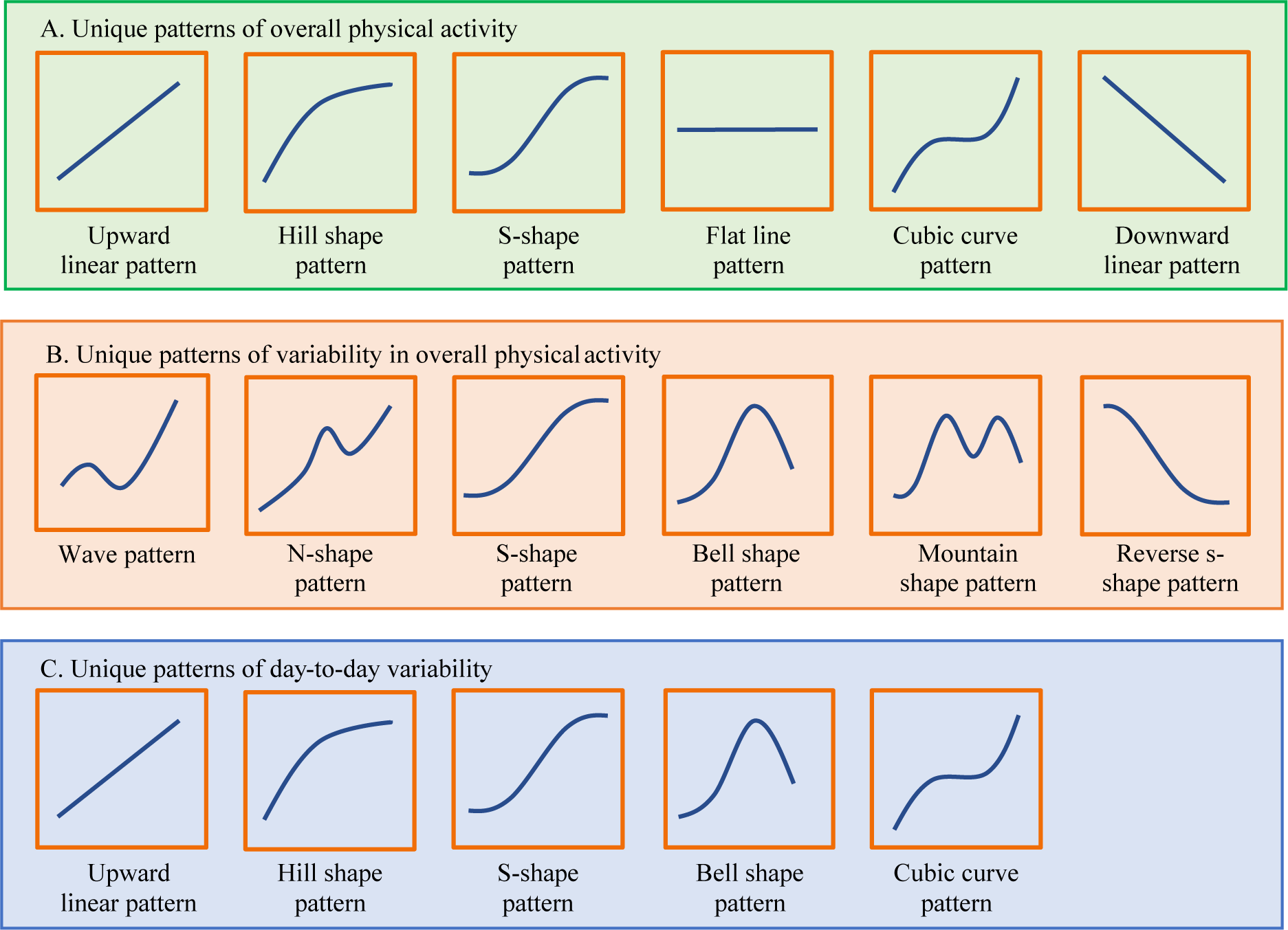
Unique patterns for each aspect of physical activity. Unique patterns were based on visual analyses and defined with the help of two experts in the geriatric rehabilitation.

For overall physical activity, six patterns were identified and named based on the morphology of the curves (Figure 3A): (1) upward linear pattern, wherein overall physical activity linearly increased during rehabilitation; (2) hill shape pattern, wherein overall physical activity initially increased but subsequently flattened; (3) S-shape pattern, wherein overall physical activity slowly increased, then steeply increased but subsequently flattened; (4) flat line pattern, wherein there was no apparent increase in overall physical activity; (5) cubic curve pattern, wherein overall physical activity initially increased, then flattened, and subsequently increased again; and (6) downward linear pattern, wherein overall physical activity gradually decreased.

For the variability in overall physical activity, six patterns were identified (Figure 3B): (1) wave pattern, wherein variability initially increased, then decreased, and subsequently increased again; (2) N-shape pattern, wherein variability initially slowly increased, then steeply increased, then decreased and lastly increased again; (3) S-shape pattern, which is similar as in the previous aspect; (4) bell-shape pattern, wherein variability initially increased but subsequently decreased; (5) mountain shape pattern, wherein variability alternately increased and decreased; and (6) reverse S-shape pattern, wherein variability slowly decreased, then steeply decreased and subsequently flattened.

For the day-to-day variability, five patterns were identified consisting of similar patterns as shown for the overall physical activity and the variability in overall physical activity (Figure 3C): (1) upward linear pattern; (2) hill shape pattern; (3) S-shape pattern; (4) bell shape pattern; and (5) cubic curve pattern.

#### Visual analysis of physical activity patterns

Once the unique patterns were identified, 18 raters independently undertook visual analysis and classified the physical activity patterns of each enrolled patient into one of the predefined unique patterns for all three aspects of physical activity. The raters were health care professionals from ZGT and the collaborating skilled nursing homes: one trauma surgeon, two physicians, two nurse practitioners, three nursing home physicians, and 10 physiotherapists with rich experience in treating older patients with a hip fracture. When a pattern did not match with any of the predefined patterns, the rater chose the option “Else”. The most frequently chosen pattern was treated as the final pattern of physical activity for each patient in all three aspects of physical activity. When there were two patterns frequently chosen for a patient, and the difference of the number of votes was equal to or smaller than two, the two experts in the geriatric rehabilitation field made the final decision of the controversial pattern.

### Statistical Analysis

To assess the reliability of the final physical activity patterns classified by the 18 raters, the interrater agreement (IRA) was estimated for all three aspects of physical activity by calculating the overall agreement between experts and the Gwet’s multirater agreement coefficient AC1.^30–33^ The level of agreement was assessed using the following classification criteria: almost perfect agreement = 0.81-1.00, substantial agreement = 0.61-0.8, moderate agreement = 0.41-0.6, fair agreement = 0.21-0.4, slight agreement = 0.00-0.2, and poor agreement = <0.00.^34^ IRA agreement analyses were performed using RStudio software (Version 1.4.1717).

Once the patterns of overall physical activity, variability in overall physical activity and day-to-day variability were known for all older patients, the overlap between the patterns was assessed. For this it was investigated whether patients with a specific pattern in one aspect of physical activity were also classified together into the same pattern for the other aspects of physical activity. When >50% of the patients have the same combination of patterns for two different aspects, these aspects were considered as overlapping and not unique aspects.

Associations between the physical activity patterns and patient characteristics were analyzed. All continuous patient variables were described as mean with the standard deviation (SD) or as median with the interquartile range (IQR) (in case of nonparametric data). All categorical variables were described as number with corresponding percentage. A Kruskal-Wallis test for continuous variables and a Fisher’s Exact Test for categorical variables were used to analyze overall differences between patterns. A p<0.05 was regarded as statistically significant. Analyses were performed using IBM SPSS statistics (version 25).

## RESULTS

A total of 82 older patients were included in this study. Of those patients, 16 were excluded due to multiple reasons, which included not completing the study (n=6), decline in health status (n=2), readmission to the hospital (n=2), COVID-19 measures (n=2), delirium (n=1), an allergic reaction to the MOX plaster (n=1), contact isolation (n=1), and complications during hospital admission (n=1). Data of the remaining 66 patients were used for further analysis. The mean (SD) age of the patients was 83.2 (6.4) years, and 49 (74.2%) patients were female. A total of 47 patients (71.2%) lived at home without help prior to the hip fracture, 17 patients (25.8%) lived at home with help, one patient (1.5%) lived at a residential home, and one patient (1.5%) lived at a nursing home. The median (IQR) duration of stay at the geriatric rehabilitation department was 28 (21-42) days. Patient characteristics are shown in Table 1.

**Table 1:** Patient characteristics

|  | **Total (n=66)** |
| --- | --- |
| **Age; median (IQR)** | 83 (79-88) |
| **Female gender; n (%)** | 49 (74.2) |
| **Premorbid living situation; n (%)** |  |
| Home | 47 (71.2) |
| Home with care | 17 (25.8) |
| Residential home | 1 (1.5) |
| Nursing home | 1 (1.5) |
| **PFMS; n (%)** |  |
| 1 | 27 (40.9) |
| 2 | 6 (9.1) |
| 3 | 33 (50) |
| 4 | - |
| 5 | - |
| **Premorbid Katz-ADL; median (IQR)** | 0 (0-1) |
| **Charlson Comorbidity Index; median (IQR)** | 1 (0-2) |
| **Surgical treatment; n (%)** |  |
| Hemiarthroplasty | 25 (37.9) |
| Intramedullary implant | 36 (54.5) |
| Dynamic Hip Screw | 5 (7.6) |
| **Weight bearing protocol admission rehabilitation; n (%)** |  |
| Non weight bearing | - |
| Partial weight bearing | 3 (4.5) |
| Full weight bearing | 63 (95.5) |
| **MoCA; median (IQR) <sup>a</sup>** | 21 (17-24) |
| **BI admission rehabilitation; median (IQR) <sup>b</sup>** | 12 (9-14) |
| **FMS admission rehabilitation; n (%)** |  |
| 1 | - |
| 2 | - |
| 3 | 7 (10.6) |
| 4 | 55 (83.3) |
| 5 | 4 (6.1) |
| **FAC score admission rehabilitation; n (%)** |  |
| 0 | 4 (6.1) |
| 1 | 4 (6.1) |
| 2 | 18 (27.3) |
| 3 | 30 (45.5) |
| 4 | 10 (15.2) |
| 5 | - |
| **Complications during rehabilitation; n (%)** |  |
| Yes | 13 (19.7) |
| No | 53 (80.3) |
| **Duration of rehabilitation stay; median (IQR)** | 28 (21-42) |
| **BI discharge rehabilitation; median (IQR) <sup>c</sup>** | 16 (15-18) |
| **FMS discharge rehabilitation; n (%) <sup>d</sup>** |  |
| 1 | - |
| 2 | 7 (10.9) |
| 3 | 42 (65.6) |
| 4 | 14 (21.9) |
| 5 | 1 (1.5) |
| **FAC score discharge rehabilitation; n (%)** |  |
| 0 | - |
| 1 | - |
| 2 | 2 (3) |
| 3 | 2 (3) |
| 4 | 51 (77.3) |
| 5 | 11 (16.7) |
| BI = Barthel Index, (P)FMS = (Pre-)Fracture Mobility Score, MoCA = Montreal Cognitive Assessment, FAC = Functional Ambulation Categories, <sup>a</sup> number of missing = 16, <sup>b</sup> number of missing = 8, <sup>c</sup> number of missing = 3, <sup>d</sup> number of missing = 2 |  |

### Physical Activity Patterns

Results of the visual analysis of the patterns of overall physical activity showed that 23 patients (34.8%) were classified with the S-shape pattern of physical activity, 15 patients (22.7%) with the upward linear pattern, six patients (9.1%) with the hill shape pattern, and six patients (9.1%) with the cubic curve pattern. The remaining patients (n=16, 24.3%) were classified as “Else”. No patients were classified with the flat line pattern or the downward linear pattern. A moderate agreement was found between the raters (overall agreement = 58% and Gwet’s AC1 = 0.52).

For the patterns of variability in overall physical activity, it was found that 14 patients (21.2%) had the N-shape pattern of variability, 10 patients (15.2%) the wave pattern, 10 patients (15.2%) the S-shape pattern, nine patients (13.6%) the bell shape pattern, nine patients (13.6%) the mountain shape pattern, and five patients (7.6%) the reversed S-shape pattern. A total of nine patients (13.6%) were classified as “Else”. A moderate agreement was found between the raters (overall agreement = 57% and Gwet’s AC1 = 0.50).

Results of the visual analysis of the patterns of day-to-day variability showed that 23 patients (34.8%) were classified with the S-shape pattern of day-to-day variability, nine patients (13.6%) with the upward linear pattern, seven patients (10.6%) with the bell shape pattern, seven patients (10.6%) with the cubic curve pattern, and six patients (9.1%) with the hill shape pattern. A total of 14 patients (21.2%) were classified as “Else”. A moderate agreement was found between the raters (overall agreement = 56% and Gwet’s AC1 = 0.48). For more details about the ratings of the independent raters see Appendix B.

### Overlap Between the Physical Activity Aspects

Results showed that the patterns of the overall physical activity and the patterns of the day-to-day variability were overlapping. All patients with the upward linear pattern of day-to-day variability also had the upward linear pattern in overall physical activity. Furthermore, four out of six patients (66.7%) with the hill shape pattern, 19 out of 23 patients (82.6%) with the S-shape pattern, and five out of seven patients (71.4%) with the cubic curve pattern had similar patterns in both aspects of physical activity (Table 2A). The aspect of the variability in overall physical activity was not overlapping with the other physical activity aspects (Table 2B-C).

**Table 2:** assessment of the overlap between the patterns of physical activity of the different aspects.

| A. overall physical activity versus day-to-day variability |  |  |  |  |  |  |
| --- | --- | --- | --- | --- | --- | --- |
| Day-to-day variability<br>Overall physical activity | Upward linear pattern | Hill shape pattern | S-shape pattern | Bell shape pattern | Cubic curve pattern | Else |
| Upward linear pattern | 9 | 0 | 3 | 0 | 1 | 0 |
| Hill shape pattern | 0 | 4 | 1 | 0 | 0 | 1 |
| S-shape pattern | 0 | 2 | 19 | 2 | 0 | 2 |
| Cubic curve pattern | 0 | 0 | 0 | 0 | 5 | 1 |
| Else | 0 | 0 | 0 | 5 | 1 | 10 |

| B. variability in overall physical activity versus day-to-day variability |  |  |  |  |  |  |
| --- | --- | --- | --- | --- | --- | --- |
| Day-to-day variability<br>Variability in overall physical activity | Upward linear pattern | Hill shape pattern | S-shape pattern | Bell shape pattern | Cubic curve pattern | Else |
| Wave pattern | 0 | 2 | 4 | 2 | 2 | 0 |
| N-shape pattern | 1 | 0 | 6 | 2 | 2 | 3 |
| S-shape pattern | 6 | 0 | 3 | 0 | 1 | 0 |
| Bell shape pattern | 1 | 0 | 5 | 2 | 0 | 1 |
| Mountain pattern | 0 | 0 | 4 | 0 | 0 | 5 |
| Reverse s-shape pattern | 0 | 3 | 0 | 1 | 1 | 0 |
| Else | 1 | 1 | 1 | 0 | 1 | 5 |

| C. variability in overall physical activity versus overall physical activity |  |  |  |  |  |
| --- | --- | --- | --- | --- | --- |
| Overall physical activity<br>Variability in overall physical activity | Upward linear pattern | Hill shape pattern | S-shape pattern | Cubic curve pattern | Else |
| Wave pattern | 0 | 2 | 5 | 2 | 1 |
| N-shape pattern | 3 | 0 | 8 | 1 | 2 |
| S-shape pattern | 7 | 1 | 1 | 1 | 0 |
| Bell shape pattern | 2 | 0 | 4 | 0 | 3 |
| Mountain pattern | 0 | 0 | 4 | 1 | 4 |
| Reverse s-shape pattern | 1 | 2 | 0 | 1 | 1 |
| Else | 2 | 1 | 1 | 0 | 5 |

### Patient Characteristics

Since the patterns of overall physical activity and the patterns of day-to-day variability were overlapping, it was chosen to exclude the day-to-day variability from further analysis and keep the overall physical activity and variability in overall physical activity. Significant associations were found between the patterns of overall physical activity and the BI at admission to the rehabilitation (BI of 15, 13, 11, 9, and 9 for, respectively, the upward linear pattern, hill shape pattern, S-shape pattern, cubic curve pattern, and “Else” group, p=0.03) and the duration of rehabilitation stay (duration of 16 days, 21 days, 30 days, 42 days, and 42 days for, respectively, the upward linear pattern, hill shape pattern, S-shape pattern, cubic curve pattern, and “Else” group, p<0.001). These significant associations, including associations that were close to statistically significant (p<0.1), are shown in Table 3. More specific, older patients with the upward linear pattern of physical activity over time or the hill shape pattern had better mobility and functionality scores at admission to the rehabilitation and a shorter duration of rehabilitation stay compared with the other patterns. The lowest complication rate was seen in older patients with the upward linear pattern or the S-shape pattern. Older patients in the “Else” group scored lowest on cognitive functioning. For the complete statistical analysis see Appendix C.

**Table 3:** Patient characteristics versus the patterns of overall physical activity.

|  | **Upward linear pattern (n=15)** | **Hill shape pattern (n=6)** | **S-shape pattern (n=23)** | **Cubic curve pattern (n=6)** | **Else (n=16)** | **p-value** |
| --- | --- | --- | --- | --- | --- | --- |
| **MoCA; median (IQR) <sup>a</sup>** | 22 (20-25) | 20 (18-21) | 21 (17-25) | 23 (21-25) | 17 (16-21) | 0.07 |
| **BI admission rehabilitation; median (IQR) <sup>b</sup>** | 15 (11-16) | 13 (12-16) | 11 (9-12) | 9 (9-11) | 9 (8-13) | **0.03** |
| **FMS admission rehabilitation; n (%)** |  |  |  |  |  | 0.06 |
| 1 | - | - | - | - | - |  |
| 2 | - | - | - | - | - |  |
| 3 | 5 (33.3) | - | - | - | 2 (12.5) |  |
| 4 | 10 (66.7) | 6 (100) | 21 (91.3) | 5 (83.3) | 13 (81.3) |  |
| 5 | - | - | 2 (8.7) | 1 (16.7) | 1 (6.3) |  |
| **FAC score admission rehab; n (%)** |  |  |  |  |  | 0.05 |
| 0 | - | - | 2 (8.7) | 1 (16.7) | 1 (6.3) |  |
| 1 | - | - | 2 (8.7) | - | 2 (12.5) |  |
| 2 | 1 (6.7) | 1 (16.7) | 7 (30.4) | 3 (50) | 6 (37.5) |  |
| 3 | 8 (53.5) | 3 (50) | 11 (47.8) | 1 (16.7) | 7 (43.8) |  |
| 4 | 6 (40) | 2 (33.3) | 1 (4.3) | 1 (16.7) | - |  |
| 5 | - | - | - | - | - |  |
| **Complications during rehabilitation; n (%)** |  |  |  |  |  | 0.07 |
| Yes | 1 (6.7) | 2 (33.3) | 2 (8.7) | 2 (33.3) | 6 (37.5) |  |
| No | 14 (93.3) | 4 (66.7) | 21 (91.3) | 4 (66.7) | 10 (62.5) |  |
| **Duration of rehabilitation stay; median (IQR)** | 16 (12-24) | 21 (10-30) | 30 (23-41) | 42 (21-47) | 42 (31-52) | **<0.001** |
| BI = Barthel Index, FMS = Fracture Mobility Score, MoCA = Montreal Cognitive Assessment, FAC = Functional Ambulation Categories, <sup>a</sup> number of missing = 16, <sup>b</sup> number of missing = 8 |  |  |  |  |  |  |

For the variability in overall physical activity, a significant association was found between the different patterns and the duration of rehabilitation stay (duration of 33 days, 35 days, 15 days, 23 days, and 42 days, 10 days, 32 days for, respectively, the wave pattern, N-shape pattern, S-shape pattern, bell shape pattern, mountain shape pattern, reverse S-shape pattern, and “Else” group, p<0.001). This significant association, including the associations that were close to statistically significant (p<0.1), are shown in Table 4. More specific, older patients with the reverse S-shape pattern had the shortest duration of rehabilitation stay followed by older patients with the S-shape pattern. Older patients with the mountain shape pattern scored lowest on cognitive functioning.

**Table 4:** Patient characteristics versus the patterns of variability in overall physical activity.

|  | **Wave**<br>**pattern**<br>**(n=10)** | **N-**<br>**shape**<br>**pattern**<br>**(n=14)** | **S-shape**<br>**pattern**<br>**(n=10)** | **Bell**<br>**shape**<br>**pattern**<br>**(n=9)** | **Mountain**<br>**shape**<br>**pattern**<br>**(n=9)** | **Reverse**<br>**S-shape**<br>**pattern**<br>**(n=5)** | **Else**<br>**(n=9)** | **p-**<br>**value** |
| --- | --- | --- | --- | --- | --- | --- | --- | --- |
| **MoCa; median**<br>**(IQR)<sup>a</sup>** | 23 (21-25) | 20 (17-24) | 23 (21-25) | 20 (17-22) | 17 (15-20) | 19 (18-20) | 21 (18-24) | 0.08 |
| **Duration of**<br>**rehabilitation**<br>**stay; median (IQR)** | 33 (21-42) | 35 (27-48) | 15 (11-25) | 23 (21-29) | 42 (41-65) | 10 (9-17) | 32 (21-42) | **<0.001** |
| MoCA = Montreal Cognitive Assessment, <sup>a</sup> number of missing =16 |  |  |  |  |  |  |  |  |

## DISCUSSION

To the best of our knowledge, this was the first study investigating continuously monitored physical activity patterns over time in older patients rehabilitating after hip fracture surgery using a MOX device. Three different aspects of physical activity were investigated: (A) the overall physical activity; (B) the variability in overall physical activity; and (C) the day-to-day variability. Results of this study showed multiple patterns for each aspect of physical activity, wherein the S-shape pattern was the most common pattern for the overall physical activity (n=23, 34.8%) and the day-to-day variability (n=23, 34.8%). The N-shape pattern was the most common pattern for the variability in overall physical activity (n=14, 21.2%). We also assessed whether there was an association between the patterns of physical activity and patient characteristics. Functionality at admission to rehabilitation and the duration of rehabilitation stay were found to be associated with the physical activity patterns of older patients during hip fracture rehabilitation.

Results of this study showed that the patterns of overall physical activity found for the enrolled patients were all increasing curves indicating that patients improved in their physical activity during rehabilitation. The S-shape pattern of overall physical activity was the most common pattern found in this study, which showed a slow increase in physical activity at the beginning of the rehabilitation. Low physical activity levels are explained by the low mobility levels of the patients, since 95.7% of the patients were not able to walk without the help of a healthcare professional at admission to rehabilitation (Table 3). According to previous literature, a slow increase in patient’s physical activity level is associated with a slow recovery in mobility and functionality. ^4, 7, 11, 12, 22, 35^ This can result in a longer duration of rehabilitation stay and a longer duration of final full functional recovery. Therefore, early mobilization and high physical activity as soon as possible during rehabilitation are highly important for a patient’s recovery. This is in line with our results showing a relationship between mobility and functionality at admission to the rehabilitation and the upward linear pattern or hill shape pattern of overall physical activity. The S-shape pattern found in this study can alert healthcare professionals of their patients’ initial slower recovery. This notification enables healthcare professionals to take action, and this may lead to more frequent mobilization of their patients after hip fracture surgery. For these patients it also might be useful to increase their confidence to be physically active without increasing the risk of falling.

Our study shows the potential of 24/7 intensity of physical activity monitoring by the fact that it gives valuable insights into older patients’ activity progress during rehabilitation. This insights into the patterns of overall physical activity could be clinically useful for multiple stakeholders. First, it could benefit healthcare organizations, since the results of this study showed that the duration of rehabilitation stay significantly differed between the patterns of overall physical activity. Older patients with an upward linear pattern had a shorter duration of rehabilitation stay than older patients with a different pattern. When the expected pattern of overall physical activity is known early in the rehabilitation, it could be used to give an indication about the duration of rehabilitation stay, which can optimize discharge planning and the patient flow within the healthcare organizations. Secondly, insights into the patterns of overall physical activity over time could be clinically useful for healthcare professionals. More information about the patterns of physical activity over time can help healthcare professionals to assess whether a patient is on track is or not. It also gives them the opportunity to adjust the rehabilitation when a patient’s progress takes a different course or when the pattern of physical activity is unfavorable. Lastly, insights into the patterns of overall physical activity over time could also help the patients themselves. Pattern information can give patients an indication of what to expect during rehabilitation, it can provide personalized feedback about the progress during rehabilitation, and it can also motivate them to be more physically active during rehabilitation. Patient motivation is considered as an important aspect of the rehabilitation. A review of Beer et al. showed that meaningful feedback about physical activity had a positive influence on the motivation of patients recovering after hip fracture surgery, which resulted in more physical activity.^36^ To further increase the clinical usefulness for all stakeholders it is considered important to know the pattern of overall physical activity over time early in the postoperative rehabilitation phase. Therefore, a recommendation for future work is to predict the patterns of overall physical activity in an early stage of postoperative rehabilitation.

Three different physical activity aspects were chosen in this study to assess physical activity in different ways. The aspect of overall physical activity was chosen, since monitoring overall physical activity directly indicates an older patient’s general activity level, which is assumed to increase during rehabilitation when older patients recover after hip fracture surgery.^4, 11^ The overall physical activity is considered as clinically relevant and easy to understand for healthcare professionals. Variability measures, on the other hand, were chosen to assess how older patients vary in their physical activity during rehabilitation. Literature showed a high within-subject variability in physical activity between days among healthy older adults, both measured in community-dwelling older adults as well as in older adults three months after hip fracture surgery.^37, 38^ For this reason, the pattern of variability in overall physical activity in older patients is expected to show an increase towards the end of rehabilitation, since we assumed that older patients were considerably recovered at the end of rehabilitation. Results of this study showed multiple patterns for the variability in overall physical activity, where five out of six patterns showed indeed an increase during rehabilitation. However, our results also showed one pattern of variability in overall physical activity with a decreasing shape. Surprisingly and without a clear explanation, patients with this pattern had the shortest duration of rehabilitation stay. More research is needed to gain more insight into the different shapes of the patterns of variability in overall physical activity.

The analyses of the physical activity patterns in this study were based on the visual analysis of multiple healthcare professionals with experience in treating older patients with a hip fracture. Multiple raters were chosen to increase the reliability of the study results, since assigning patterns to a patient’s overall physical activity progress can be subject to subjectivity. IRA analysis showed a moderate agreement between the raters for all three physical activity aspects, which indicates that we currently cannot perform visual pattern analyses based on one rater. Two potential reasons for moderate agreement found in this study are discussed here: (1) the high number of patterns for each aspect made a high IRA difficult to reach.^39^ In this study, we summarized six different patterns of overall physical activity given the large heterogeneity of the older patients’ progress of physical activity during rehabilitation. The high number of patterns could cause the moderate IRA, but more importantly these patterns can comprehensively cover the different overall physical activity patterns of older patients after hip fracture surgery. (2) We deliberately chose to not train the raters to ensure the independence of raters during the visual analysis in this study. The raters without relevant training given by an expert may give different pattern judgement based on their own clinical practice experiences with older patients. Such subjectivity could influence the IRA, but the moderate IRA found in our study exactly reflected the nature of clinical judgement that we cannot avoid. Consequently, we handled the moderate IRA among the raters through organizing an expert meeting with two experts in the geriatric rehabilitation field to make a final decision on the patterns of the questionable patients. In this way, we guaranteed the reliability of our study.

This study also had some limitations. The first limitation was the low number of enrolled patients (n=66) making this study a preliminary study. Due to this low number, the statistics were kept more descriptive and only assessed the overall differences between the patient variables and the patterns of physical activity for each aspect of physical activity. Statistical results from this study should, therefore, be interpreted cautiously and multivariate analysis to assess which patient variables are meaningful to differentiate between physical activity patterns could not be performed. A second limitation was that not all enrolled patients could be classified in one of the pre-defined patterns of overall physical activity over time. More research is recommended to also classify or predict the patterns of the older patients in the “Else” group. More specific classification and prediction of the patterns currently classified as “Else” could further increase the clinical usefulness of assessing the patterns of physical activity in older patients after hip fracture surgery. A final limitation was that we did not include pain and fear of falling as parameters in this study. Literature shows that high fear of falling, and pain after hip fracture surgery have a negative effect on the physical activity of older patients, which could also affect the pattern of overall physical activity over time.^9, 11, 12, 40, 41^ More research is recommended to also assess the influence of fear of falling and pain on the patterns of overall physical activity over time.

## CONCLUSION

The aim of this preliminary study was to investigate the patterns of physical activity in older patients rehabilitating after hip fracture surgery and to assess the association between the patterns of physical activity and patient characteristics. This study showed different patterns of physical activity during rehabilitation, in which it was shown that most patients had a positive evolution in their physical activity over time during rehabilitation after hip fracture surgery. The S-shape pattern was the most common pattern for the overall physical activity and the day-to-day variability. The N-shape was the most common for the variability in overall physical activity. Functionality at admission to the rehabilitation, measured with the BI, was associated with the patterns of physical activity. Furthermore, a significant difference in the duration of geriatric rehabilitation stay was found between the patterns of physical activity. The difference found in physical activity patterns in older patients during hip fracture rehabilitation could be relevant for healthcare organization, healthcare professionals and patients to optimize and personalize hip fracture treatment.

## Data Availability

All data produced in the present study are not available

## Appendix A. Workflow data-analysis raw MOX data

| Steps for analysing raw MOX data |  | Remarks |
| --- | --- | --- |
| 1. Pre-processing | Select data between 7.00 am and 10 pm<br>Detection of missing data:<br>More than 10 missing minutes in one hour? Yes: define hour as missing hour<br>More than 3 missing hours in one day? Yes: define day as a missing day<br>For the remaining days:<br>Moving average filter with a 0.12 second window size<br>High pass filter with a cut-off frequency of 1 Hz<br>Data segmentation with a window size of 2 seconds. | Missing data is detected in the raw data by checking for missing timestamps.<br>A missing day was classified with a NaN value for the intensity of physical activity.<br>Removal of noise acceleration and gravitational acceleration using a moving average filter and high pass filter <sup>42</sup>.<br>Data segmentation results in 27,000 windows per day. |
| 2. Calculate the physical activity parameter for each physical activity aspect | Intensity of physical activity:<br>Calculate the Signal Magnitude Area (SMA) for each window of 2 seconds.<br>Take the sum of the SMA of all windows in a day to calculate the overall intensity of physical activity per day (counts/day)<br>Variability in the intensity of physical activity:<br>Data segmentation with a window size of 5 days<br>Calculate the variability for each window with the “*var*” function in Matlab<br>Variability in the intensity of physical activity within each day:<br>Take the SMA of all windows in a day (27.000 windows)<br>Calculate the variability for each day (over all 27.000 windows) with the “*var*” function in Matlab | For the variability in the intensity of physical activity an overlapping, centred sliding window method was used. |
| 4. Plot figures | Smooth the data with a Gaussian-weighted moving average smoothing filter with a window length of 14 days using the function “*smoothdata*” in Matlab<br>Plot the overall intensity of physical activity and the day-to-day variability over time<br>Plot the variability in overall intensity of physical activity for each window | The smoothing filter used an overlapping centred sliding window method. |

## Appendix B. Ratings for each patient by all raters and for each aspect of physical activity

Appendix B. Ratings for each patient by all raters and for each aspect of physical activity A. Pattern of overall physical activity
| Enrolled patients | Upward linear pattern | Hill shape pattern | S-shape pattern | Flat line pattern | Cubic curve pattern | Downward linear pattern | Else |
| --- | --- | --- | --- | --- | --- | --- | --- |
| 1 | 0 | 3 | 13 | 0 | 2 | 0 | 0 |
| 2 | 0 | 0 | 1 | 2 | 0 | 1 | 14 |
| 3 | 0 | 0 | 3 | 0 | 15 | 0 | 0 |
| 4 | 0 | 3 | 1 | 2 | 0 | 0 | 12 |
| 5 | 1 | 0 | 5 | 0 | 2 | 0 | 10 |
| 6 | 3 | 11 | 4 | 0 | 0 | 0 | 0 |
| 7 | 0 | 0 | 1 | 0 | 15 | 0 | 2 |
| 8 | 5 | 1 | 7 | 0 | 4 | 0 | 1 |
| 9 | 2 | 6 | 10 | 0 | 0 | 0 | 0 |
| 10 | 0 | 1 | 2 | 0 | 15 | 0 | 0 |
| 11 | 1 | 0 | 2 | 1 | 4 | 0 | 10 |
| 12 | 18 | 0 | 0 | 0 | 0 | 0 | 0 |
| 13 | 1 | 1 | 6 | 0 | 2 | 0 | 8 |
| 14 | 0 | 0 | 0 | 4 | 0 | 3 | 11 |
| 15 | 1 | 0 | 0 | 6 | 0 | 0 | 11 |
| 16 | 15 | 1 | 2 | 0 | 0 | 0 | 0 |
| 17 | 0 | 1 | 0 | 2 | 0 | 0 | 15 |
| 18 | 3 | 0 | 2 | 0 | 13 | 0 | 0 |
| 19 | 0 | 0 | 14 | 0 | 3 | 0 | 1 |
| 20 | 0 | 0 | 0 | 1 | 6 | 0 | 11 |
| 21 | 4 | 0 | 1 | 1 | 5 | 0 | 7 |
| 22 | 2 | 0 | 12 | 0 | 2 | 0 | 2 |
| 23 | 4 | 2 | 12 | 0 | 0 | 0 | 0 |
| 24 | 18 | 0 | 0 | 0 | 0 | 0 | 0 |
| 25 | 18 | 0 | 0 | 0 | 0 | 0 | 0 |
| 26 | 0 | 1 | 17 | 0 | 0 | 0 | 0 |
| 27 | 10 | 0 | 6 | 0 | 2 | 0 | 0 |
| 28 | 18 | 0 | 0 | 0 | 0 | 0 | 0 |
| 29 | 12 | 5 | 1 | 0 | 0 | 0 | 0 |
| 30 | 0 | 0 | 0 | 2 | 3 | 0 | 13 |
| 31 | 7 | 0 | 5 | 0 | 1 | 0 | 5 |
| 32 | 0 | 0 | 18 | 0 | 0 | 0 | 0 |
| 33 | 1 | 2 | 8 | 0 | 4 | 0 | 3 |
| 34 | 17 | 0 | 1 | 0 | 0 | 0 | 0 |
| 35 | 0 | 5 | 10 | 0 | 0 | 0 | 3 |
| 36 | 1 | 0 | 16 | 0 | 0 | 0 | 1 |
| 37 | 0 | 1 | 17 | 0 | 0 | 0 | 0 |
| 38 | 1 | 2 | 15 | 0 | 0 | 0 | 0 |
| 39 | 1 | 2 | 3 | 0 | 3 | 0 | 9 |
| 40 | 0 | 1 | 4 | 0 | 4 | 0 | 9 |
| 41 | 3 | 13 | 2 | 0 | 0 | 0 | 0 |
| 42 | 4 | 1 | 2 | 0 | 11 | 0 | 0 |
| 43 | 7 | 0 | 6 | 0 | 2 | 0 | 3 |
| 44 | 0 | 0 | 13 | 0 | 1 | 0 | 4 |
| 45 | 0 | 0 | 0 | 6 | 0 | 0 | 12 |
| 46 | 0 | 1 | 17 | 0 | 0 | 0 | 0 |
| 47 | 1 | 0 | 15 | 0 | 0 | 0 | 2 |
| 48 | 0 | 0 | 13 | 0 | 1 | 0 | 4 |
| 49 | 16 | 1 | 1 | 0 | 0 | 0 | 0 |
| 50 | 16 | 1 | 0 | 1 | 0 | 0 | 0 |
| 51 | 16 | 0 | 2 | 0 | 0 | 0 | 0 |
| 52 | 2 | 7 | 9 | 0 | 0 | 0 | 0 |
| 53 | 0 | 0 | 15 | 0 | 0 | 0 | 3 |
| 54 | 0 | 1 | 0 | 2 | 2 | 1 | 12 |
| 55 | 4 | 1 | 7 | 0 | 3 | 0 | 3 |
| 56 | 1 | 0 | 15 | 0 | 0 | 0 | 2 |
| 57 | 0 | 0 | 16 | 0 | 1 | 0 | 1 |
| 58 | 5 | 11 | 1 | 0 | 1 | 0 | 0 |
| 59 | 0 | 6 | 0 | 0 | 7 | 0 | 5 |
| 60 | 17 | 0 | 1 | 0 | 0 | 0 | 0 |
| 61 | 3 | 15 | 0 | 0 | 0 | 0 | 0 |
| 62 | 0 | 17 | 0 | 0 | 0 | 0 | 1 |
| 63 | 2 | 0 | 0 | 0 | 15 | 0 | 1 |
| 64 | 0 | 4 | 0 | 1 | 0 | 1 | 12 |
| 65 | 0 | 1 | 1 | 0 | 5 | 0 | 11 |
| 66 | 0 | 2 | 16 | 0 | 0 | 0 | 0 |

B. Pattern of variability in overall physical activity
| Enrolled patients | Wave pattern | N-shape pattern | S-shape pattern | Bell shape pattern | Mountain pattern | Reverse s-shape pattern | Else |
| --- | --- | --- | --- | --- | --- | --- | --- |
| 1 | 0 | 6 | 1 | 1 | 5 | 0 | 5 |
| 2 | 0 | 0 | 0 | 17 | 0 | 0 | 1 |
| 3 | 2 | 0 | 11 | 0 | 0 | 0 | 5 |
| 4 | 0 | 0 | 0 | 18 | 0 | 0 | 0 |
| 5 | 1 | 0 | 0 | 8 | 4 | 0 | 5 |
| 6 | 8 | 1 | 6 | 1 | 0 | 0 | 2 |
| 7 | 0 | 0 | 0 | 0 | 11 | 0 | 7 |
| 8 | 0 | 0 | 0 | 18 | 0 | 0 | 0 |
| 9 | 0 | 0 | 0 | 15 | 0 | 0 | 3 |
| 10 | 0 | 0 | 0 | 0 | 0 | 12 | 6 |
| 11 | 13 | 0 | 3 | 0 | 0 | 0 | 2 |
| 12 | 0 | 0 | 17 | 0 | 0 | 0 | 1 |
| 13 | 1 | 1 | 1 | 6 | 0 | 0 | 9 |
| 14 | 0 | 7 | 0 | 6 | 4 | 0 | 1 |
| 15 | 2 | 1 | 0 | 0 | 1 | 0 | 14 |
| 16 | 0 | 0 | 0 | 14 | 0 | 2 | 2 |
| 17 | 3 | 0 | 1 | 0 | 0 | 0 | 14 |
| 18 | 17 | 1 | 0 | 0 | 0 | 0 | 0 |
| 19 | 5 | 2 | 6 | 1 | 0 | 0 | 4 |
| 20 | 0 | 3 | 0 | 5 | 2 | 0 | 8 |
| 21 | 0 | 2 | 0 | 0 | 11 | 0 | 5 |
| 22 | 1 | 0 | 15 | 0 | 0 | 0 | 2 |
| 23 | 0 | 0 | 0 | 0 | 14 | 0 | 4 |
| 24 | 7 | 0 | 9 | 0 | 0 | 0 | 2 |
| 25 | 0 | 0 | 18 | 0 | 0 | 0 | 0 |
| 26 | 7 | 1 | 7 | 0 | 0 | 0 | 3 |
| 27 | 0 | 0 | 15 | 0 | 0 | 0 | 3 |
| 28 | 0 | 0 | 16 | 0 | 0 | 0 | 2 |
| 29 | 5 | 3 | 0 | 0 | 3 | 0 | 7 |
| 30 | 1 | 0 | 0 | 0 | 1 | 0 | 16 |
| 31 | 3 | 0 | 8 | 0 | 0 | 0 | 7 |
| 32 | 15 | 3 | 0 | 0 | 0 | 0 | 0 |
| 33 | 8 | 7 | 0 | 0 | 0 | 0 | 3 |
| 34 | 0 | 18 | 0 | 0 | 0 | 0 | 0 |
| 35 | 11 | 5 | 0 | 0 | 0 | 0 | 2 |
| 36 | 1 | 3 | 0 | 0 | 9 | 0 | 5 |
| 37 | 0 | 13 | 0 | 2 | 0 | 0 | 3 |
| 38 | 0 | 0 | 0 | 18 | 0 | 0 | 0 |
| 39 | 0 | 6 | 0 | 0 | 4 | 0 | 8 |
| 40 | 0 | 0 | 0 | 0 | 18 | 0 | 0 |
| 41 | 0 | 0 | 0 | 0 | 0 | 12 | 6 |
| 42 | 11 | 1 | 2 | 0 | 0 | 0 | 4 |
| 43 | 0 | 10 | 0 | 1 | 5 | 0 | 2 |
| 44 | 0 | 10 | 1 | 2 | 0 | 0 | 5 |
| 45 | 4 | 5 | 0 | 0 | 1 | 0 | 8 |
| 46 | 0 | 0 | 0 | 5 | 11 | 0 | 2 |
| 47 | 1 | 9 | 0 | 6 | 0 | 0 | 2 |
| 48 | 0 | 0 | 0 | 0 | 11 | 0 | 7 |
| 49 | 0 | 18 | 0 | 0 | 0 | 0 | 0 |
| 50 | 0 | 0 | 0 | 0 | 0 | 13 | 5 |
| 51 | 0 | 0 | 9 | 6 | 0 | 0 | 3 |
| 52 | 0 | 12 | 0 | 3 | 1 | 0 | 2 |
| 53 | 0 | 0 | 2 | 15 | 0 | 0 | 1 |
| 54 | 1 | 1 | 0 | 0 | 10 | 0 | 6 |
| 55 | 1 | 16 | 0 | 0 | 1 | 0 | 0 |
| 56 | 0 | 0 | 0 | 18 | 0 | 0 | 0 |
| 57 | 0 | 9 | 2 | 0 | 3 | 0 | 4 |
| 58 | 17 | 0 | 1 | 0 | 0 | 0 | 0 |
| 59 | 2 | 1 | 0 | 0 | 4 | 0 | 11 |
| 60 | 0 | 0 | 14 | 0 | 0 | 0 | 4 |
| 61 | 0 | 0 | 0 | 0 | 0 | 17 | 1 |
| 62 | 14 | 0 | 0 | 0 | 0 | 0 | 4 |
| 63 | 5 | 8 | 0 | 1 | 0 | 0 | 4 |
| 64 | 0 | 0 | 0 | 0 | 0 | 18 | 0 |
| 65 | 0 | 0 | 0 | 0 | 11 | 0 | 7 |
| 66 | 2 | 16 | 0 | 0 | 0 | 0 | 0 |

C. Pattern of day-to-day variability
| Enrolled patients | Upward linear pattern | Hill pattern | S-shape pattern | Bell shape pattern | Cubic curve pattern | Else |
| --- | --- | --- | --- | --- | --- | --- |
| 1 | 0 | 5 | 1 | 0 | 4 | 8 |
| 2 | 0 | 0 | 0 | 11 | 0 | 7 |
| 3 | 4 | 0 | 2 | 0 | 11 | 1 |
| 4 | 0 | 0 | 0 | 14 | 0 | 4 |
| 5 | 0 | 0 | 1 | 0 | 1 | 16 |
| 6 | 1 | 4 | 12 | 0 | 1 | 0 |
| 7 | 0 | 0 | 1 | 0 | 9 | 8 |
| 8 | 14 | 0 | 2 | 0 | 0 | 2 |
| 9 | 0 | 6 | 12 | 0 | 0 | 0 |
| 10 | 0 | 1 | 2 | 0 | 14 | 1 |
| 11 | 0 | 0 | 1 | 10 | 1 | 6 |
| 12 | 16 | 2 | 0 | 0 | 0 | 0 |
| 13 | 0 | 0 | 13 | 0 | 1 | 4 |
| 14 | 0 | 0 | 0 | 4 | 1 | 13 |
| 15 | 0 | 1 | 6 | 2 | 0 | 9 |
| 16 | 5 | 2 | 11 | 0 | 0 | 0 |
| 17 | 0 | 0 | 0 | 1 | 3 | 14 |
| 18 | 0 | 0 | 1 | 0 | 10 | 7 |
| 19 | 0 | 0 | 11 | 1 | 3 | 3 |
| 20 | 0 | 0 | 0 | 0 | 8 | 10 |
| 21 | 0 | 0 | 6 | 0 | 3 | 9 |
| 22 | 0 | 0 | 11 | 0 | 2 | 5 |
| 23 | 0 | 1 | 14 | 1 | 1 | 1 |
| 24 | 18 | 0 | 0 | 0 | 0 | 0 |
| 25 | 18 | 0 | 0 | 0 | 0 | 0 |
| 26 | 0 | 0 | 17 | 1 | 0 | 0 |
| 27 | 1 | 1 | 15 | 0 | 0 | 1 |
| 28 | 18 | 0 | 0 | 0 | 0 | 0 |
| 29 | 5 | 8 | 5 | 0 | 0 | 0 |
| 30 | 0 | 0 | 0 | 3 | 2 | 13 |
| 31 | 16 | 0 | 1 | 0 | 1 | 0 |
| 32 | 2 | 1 | 15 | 0 | 0 | 0 |
| 33 | 0 | 0 | 9 | 0 | 4 | 5 |
| 34 | 0 | 1 | 1 | 0 | 11 | 5 |
| 35 | 0 | 3 | 2 | 8 | 0 | 5 |
| 36 | 1 | 0 | 12 | 0 | 2 | 3 |
| 37 | 0 | 1 | 7 | 9 | 0 | 1 |
| 38 | 0 | 2 | 14 | 1 | 0 | 1 |
| 39 | 0 | 0 | 2 | 8 | 0 | 8 |
| 40 | 0 | 0 | 6 | 0 | 3 | 9 |
| 41 | 0 | 15 | 2 | 0 | 0 | 1 |
| 42 | 3 | 0 | 3 | 0 | 9 | 3 |
| 43 | 0 | 1 | 7 | 0 | 6 | 4 |
| 44 | 0 | 0 | 11 | 0 | 1 | 6 |
| 45 | 0 | 0 | 0 | 5 | 0 | 13 |
| 46 | 0 | 1 | 17 | 0 | 0 | 0 |
| 47 | 0 | 1 | 15 | 2 | 0 | 0 |
| 48 | 0 | 0 | 15 | 2 | 0 | 1 |
| 49 | 9 | 4 | 5 | 0 | 0 | 0 |
| 50 | 4 | 14 | 0 | 0 | 0 | 0 |
| 51 | 10 | 0 | 7 | 0 | 0 | 1 |
| 52 | 0 | 2 | 16 | 0 | 0 | 0 |
| 53 | 0 | 0 | 11 | 0 | 2 | 5 |
| 54 | 0 | 0 | 0 | 8 | 1 | 9 |
| 55 | 0 | 0 | 0 | 0 | 4 | 14 |
| 56 | 0 | 1 | 16 | 0 | 0 | 1 |
| 57 | 0 | 1 | 15 | 0 | 0 | 2 |
| 58 | 0 | 13 | 5 | 0 | 0 | 0 |
| 59 | 0 | 3 | 1 | 0 | 7 | 7 |
| 60 | 15 | 0 | 1 | 0 | 1 | 1 |
| 61 | 1 | 17 | 0 | 0 | 0 | 0 |
| 62 | 0 | 16 | 2 | 0 | 0 | 0 |
| 63 | 0 | 1 | 1 | 0 | 12 | 4 |
| 64 | 0 | 1 | 0 | 17 | 0 | 0 |
| 65 | 0 | 1 | 0 | 6 | 1 | 10 |
| 66 | 0 | 1 | 17 | 0 | 0 | 0 |

## Appendix C. Patient variables for each pattern of physical activity of all three aspects of physical activity

Appendix C. Patient variables for each pattern of physical activity of all three aspects of physical activity A. Pattern of overall physical activity
|  | Total<br>(n=66) | Upward linear<br>pattern (n=15) | Hill shape<br>pattern (n=6) | S-shape<br>pattern (n=23) | Cubic curve<br>pattern (n=6) | Else<br>(n=16) | p-value |
| --- | --- | --- | --- | --- | --- | --- | --- |
| **Age; median (IQR)** | 83 (79-88) | 82 (81-87) | 84 (79-89) | 83 (80-90) | 83 (78-84) | 83 (78-88) | 0.90 |
| **Female gender; n (%)** | 49 (74.2) | 12 (80) | 4 (66.7) | 15 (65.2) | 5 (83.3) | 13 (81.3) | 0.79 |
| **Premorbid living situation; n (%)** |  |  |  |  |  |  |  |
| Home | 47 (71.2) | 10 (66.6) | 5 (83.3) | 16 (69.6) | 6 (100) | 10 (62.5) | 0.64 |
| Home with care | 17 (25.8) | 4 (26.7) | 1 (16.7) | 7 (30.4) | - | 5 (31.3) |  |
| Residential home | 1 (1.5) | 1 (6.7) | - | - | - | - |  |
| Nursing home | 1 (1.5) | - | - | - | - | 1 (6.3) |  |
| **PFMS; n (%)** |  |  |  |  |  |  |  |
| 1 | 27 (40.9) | 9 (60) | 2 (33.3) | 10 (43.5) | 2 (33.3) | 4 (25) | 0.69 |
| 2 | 6 (9.1) | 1 (6.7) | - | 2 (8.7) | 1 (16.7) | 2 (12.5) |  |
| 3 | 33 (50) | 5 (33.3) | 4 (66.7) | 11 (47.8) | 3 (50) | 10 (62.5) |  |
| 4 | - | - | - | - | - | - |  |
| 5 | - | - | - | - | - | - |  |
| **Premorbid Katz-ADL; median (IQR)** | 0 (0-1) | 0 (0-0) | 0 (0-0) | 0 (0-1) | 0 (0-0) | 0 (0-1) | 0.17 |
| **Charlson Comorbidity Index; median (IQR)** | 1 (0-2) | 0 (0-3) | 1 (0-1) | 1 (0-2) | 0 (0-1) | 1 (1-2) | 0.37 |
| **Surgical treatment; n (%)** |  |  |  |  |  |  |  |
| Hemiarthroplasty | 25 (37.9) | 6 (40) | 3 (50) | 8 (34.8) | 1 (16.7) | 7 (43.8) | 0.80 |
| Intramedullary implant | 36 (54.5) | 7 (46.7) | 3 (50) | 13 (56.5) | 4 (66.7) | 9 (56.3) |  |
| Dynamic Hip Screw | 5 (7.6) | 2 (13.3) | - | 2 (8.7) | 1 (16.7) | - |  |
| **Weight bearing protocol; n (%)** |  |  |  |  |  |  |  |
| Non weight bearing | - | - | - | - | - | - | 0.29 |
| Partial weight bearing | 3 (4.5) | - | - | 2 (8.7) | 1 (16.7) | - |  |
| Full weight bearing | 63 (95.5) | 15 (100) | 6 (100) | 21 (91.3) | 5 (83.3) | 16 (100) |  |
| **MoCA; median (IQR) <sup>a</sup>** | 21 (17-24) | 22 (20-25) | 20 (18-21) | 21 (17-25) | 23 (21-25) | 17 (16-21) | 0.07 |
| **BI admission rehabilitation; median (IQR) <sup>b</sup>** | 12 (9-14) | 15 (11-16) | 13 (12-16) | 11 (9-12) | 9 (9-11) | 9 (8-13) | **0.03** |
| **FMS admission rehabilitation; n (%)** |  |  |  |  |  |  |  |
| 1 | - | - | - | - | - | - | 0.06 |
| 2 | - | - | - | - | - | - |  |
| 3 | 7 (10.6) | 5 (33.3) | - | - | - | 2 (12.5) |  |
| 4 | 55 (83.3) | 10 (66.7) | 6 (100) | 21 (91.3) | 5 (83.3) | 13 (81.3) |  |
| 5 | 4 (6.1) | - | - | 2 (8.7) | 1 (16.7) | 1 (6.3) |  |
| **FAC score admission rehab; n (%)** |  |  |  |  |  |  |  |
| 0 | 4 (6.1) | - | - | 2 (8.7) | 1 (16.7) | 1 (6.3) | 0.05 |
| 1 | 4 (6.1) | - | - | 2 (8.7) | - | 2 (12.5) |  |
| 2 | 18 (27.3) | 1 (6.7) | 1 (16.7) | 7 (30.4) | 3 (50) | 6 (37.5) |  |
| 3 | 30 (45.4) | 8 (53.5) | 3 (50) | 11 (47.8) | 1 (16.7) | 7 (43.8) |  |
| 4 | 10 (15.1) | 6 (40) | 2 (33.3) | 1 (4.3) | 1 (16.7) | - |  |
| 5 | - | - | - | - | - | - |  |
| **Complications during rehabilitation; n (%)** |  |  |  |  |  |  |  |
| Yes | 13 (19.7) | 1 (6.7) | 2 (33.3) | 2 (8.7) | 2 (33.3) | 6 (37.5) | 0.07 |
| No | 53 (80.3) | 14 (93.3) | 4 (66.7) | 21 (91.3) | 4 (66.7) | 10 (62.5) |  |
| **Duration of rehabilitation stay; median (IQR)** | 28 (21-42) | 16 (12-24) | 21 (10-30) | 30 (23-41) | 42 (21-47) | 42 (31-52) | <0.001 |
| **BI discharge rehabilitation; median (IQR) <sup>c</sup>** | 16 (15-18) | 17 (17-18) | 17 (16-18) | 16 (15-18) | 17 (15-18) | 16 (14-17) | 0.21 |
| **FMS discharge rehabilitation; n (%) <sup>d</sup>** |  |  |  |  |  |  |  |
| 1 | - | - | - | - | - | - | 0.24 |
| 2 | 7 (10.9) | 3 (20) | - | 3 (13.6) | 1 (16.7) | - |  |
| 3 | 42 (65.6) | 11 (73.3) | 5 (83.3) | 13 (59.1) | 5 (83.3) | 8 (53.3) |  |
| 4 | 14 (21.9) | 1 (6.7) | 1 (16.7) | 6 (27.3) | - | 6 (40) |  |
| 5 | 1 (1.5) | - | - | - | - | 1 (6.7) |  |
| **FAC score discharge rehabilitation; n (%)** |  |  |  |  |  |  |  |
| 0 | - | - | - | - | - | - | 0.19 |
| 1 | - | - | - | - | - | - |  |
| 2 | 2 (3) | - | - | - | - | 2 (12.5) |  |
| 3 | 2 (3) | - | - | - | - | 2 (12.5) |  |
| 4 | 51 (77.3) | 10 (66.7) | 5 (83.3) | 21 (91.3) | 5 (83.3) | 10 (62.5) |  |
| 5 | 11 (16.7) | 5 (33.3) | 1 (16.7) | 2 (8.7) | 1 (16.7) | 2 (12.5) |  |
| BI = Barthel Index, (P)FMS = (Pre-)Fracture Mobility Score, MoCA = Montreal Cognitive Assessment, FAC = Functional Ambulation Categories |  |  |  |  |  |  |  |
| <sup>a</sup> number of missing = 16, <sup>b</sup> number of missing = 8, <sup>c</sup> number of missing = 3, <sup>d</sup> number of missing = 2 |  |  |  |  |  |  |  |

B. Pattern of variability in overall physical activity
|  | **Total**<br>**(n=66)** | **Wave**<br>**pattern**<br>**(n=10)** | **N-shape**<br>**pattern**<br>**(n=141)** | **S-shape**<br>**pattern**<br>**(n=10)** | **Bell**<br>**shape**<br>**pattern**<br>**(n=9)** | **Mountain**<br>**shape**<br>**pattern**<br>**(n=9)** | **Reverse**<br>**s-shape**<br>**pattern**<br>**(n=5)** | **Else**<br>**(n=9)** | **p-**<br>**value** |
| --- | --- | --- | --- | --- | --- | --- | --- | --- | --- |
| **Age; median (IQR)** | 83 (79-88) | 83 (82-89) | 84 (82-87) | 81 (74-86) | 84 (79-90) | 82 (75-85) | 83 (83-84) | 80 (79-88) | 0.83 |
| **Female gender; n (%)** | 49 (74.2) | 6 (60) | 13 (92.9) | 9 (90) | 4 (44.4) | 6 (66.7) | 4 (80) | 7 (77.8) | 0.14 |
| **Premorbid living situation; n (%)** |  |  |  |  |  |  |  |  |  |
| Home | 47 (71.2) | 8 (80) | 11 (78.6) | 6 (60) | 6 (66.7) | 6 (66.7) | 4 (80) | 6 (66.7) | 0.64 |
| Home with care | 17 (25.8) | 2 (20) | 3 (21.4) | 4 (40) | 3 (33.3) | 3 (33.3) | - | 2 (22.2) |  |
| Residential home | 1 (1.5) | - | - | - | - | - | 1 (20) | - |  |
| Nursing home | 1 (1.5) | - | - | - | - | - | - | 1 (11.1) |  |
| **PFMS; n (%)** |  |  |  |  |  |  |  |  |  |
| 1 | 27 (40.9) | 2 (20) | 7 (50) | 4 (40) | 4 (44.4) | 4 (44.4) | 2 (40) | 4 (44.4) | 0.43 |
| 2 | 6 (9.1) | 1 (10) | - | 1 (10) | 3 (33.3) | - | 1 (20) | - |  |
| 3 | 33 (50) | 7 (70) | 7 (50) | 5 (50) | 2 (22.3) | 5 (55.6) | 2 (40) | 5 (55.6) |  |
| 4 | - | - | - | - | - | - | - | - |  |
| 5 | - | - | - | - | - | - | - | - |  |
| **Premorbid Katz-ADL; median (IQR)** | 0 (0-1) | 0 (0-1) | 0 (0-0) | 0 (0-1) | 0 (0-1) | 0 (0-1) | 0 (0-0) | 0 (0-1) | 0.56 |
| **Charlson Comorbidity Index; median (IQR)** | 1 (0-2) | 1 (1-2) | 1 (0-2) | 0 (0-3) | 1 (0-3) | 1 (0-1) | 1 (0-1) | 1 (1-2) | 0.65 |
| **Surgical treatment; n (%)** |  |  |  |  |  |  |  |  |  |
| Hemiarthroplasty | 25 (37.9) | 4 (40) | 5 (35.7) | 3 (30) | 6 (66.7) | 2 (22.2) | 3 (60) | 2 (22.2) | 0.36 |
| Intramedullary implant | 36 (54.5) | 6 (60) | 9 (64.3) | 5 (50) | 3 (33.3) | 6 (66.7) | 1 (20) | 6 (66.7) |  |
| Dynamic Hip Screw | 5 (7.6) | - | - | 2 (20) | - | 1 (11.1) | 1 (20) | 1 (11.1) |  |
| **Weight bearing protocol; n (%)** |  |  |  |  |  |  |  |  |  |
| Non weight bearing | - | - | - | - | - | - | - | - | 0.58 |
| Partial weight bearing | 3 (4.5) | - | - | 1 (10) | - | 1 (11.1) | - | 1 (11.1) |  |
| Full weight bearing | 63 (95.5) | 10(100) | 14 (100) | 9 (90) | 9 (100) | 8 (88.9) | 5 (100) | 8 (88.9) |  |
| **MoCa; median (IQR) <sup>a</sup>** | 21 (17-24) | 23 (21-25) | 20 (17-24) | 23 (21-25) | 20 (17-22) | 17 (15-20) | 19 (18-20) | 21 (18-24) | 0.08 |
| **BI admission rehabilitation; median (IQR) <sup>b</sup>** | 12 (9-14) | 12 (10-14) | 10 (8-11) | 13 (12-16) | 12 (10-15) | 9 (8-12) | 14 (12-16) | 11 (9-14) | 0.17 |
| **FMS admission rehabilitation;** |  |  |  |  |  |  |  |  |  |
| 1 | - | - | - | - | - | - | - | - | 0.49 |
| 2 | - | - | - | - | - | - | - | - |  |
| 3 | 7 (10.6) | - | - | 2 (20) | 2 (22.2) | 1 (11.1) | - | 2 (22.2) |  |
| 4 | 55 (83.3) | 10 (100) | 13 (92.9) | 7 (70) | 7 (77.8) | 7 (77.8) | 5 (100) | 6 (66.7) |  |
| 5 | 4 (6.1) | - | 1 (7.1) | 1 (10) | - | 1 (11.1) | - | 1 (11.1) |  |
| **FAC score admission rehabilitation; n (%)** |  |  |  |  |  |  |  |  |  |
| 0 | 4 (6.1) | - | 1 (7.1) | 1 (10) | - | 1 (11.1) | - | 1 (11.1) | 0.29 |
| 1 | 4 (6.1) | 1 (10) | - | - | - | 2 (22.2) | - | 1 (11.1) |  |
| 2 | 18 (27.3) | 5 (50) | 5 (35.7) | 1 (10) | 3 (33.3) | 3 (33.3) | - | 1 (11.1) |  |
| 3 | 30 (45.4) | 4 (40) | 7 (50) | 5 (50) | 4 (44.5) | 3 (33.3) | 2 (40) | 5 (55.6) |  |
| 4 | 10 (15.1) | - | 1 (7.1) | 3 (30) | 2 (22.2) | - | 3 (60) | 1 (11.1) |  |
| 5 | - | - | - | - | - | - | - | - |  |
| **Complications during rehabilitation; n (%)** |  |  |  |  |  |  |  |  |  |
| Yes | 13 (19.7) | - | 3 (21.4) | 2 (20) | 3 (33.3) | 2 (22.2) | 1 (20) | 2 (22.2) | 0.67 |
| No | 53 (80.3) | 10 (100) | 11 (78.6) | 8 (80) | 6 (66.7) | 7 (77.8) | 4 (80) | 7 (77.8) |  |
| **Duration of rehabilitation stay; median (IQR)** | 28 (21-42) | 33 (21-42) | 35 (27-48) | 15 (11-25) | 23 (21-29) | 42 (41-65) | 10 (9-17) | 32 (21-42) | <0.001 |
| **BI discharge rehabilitation; median (IQR) <sup>c</sup>** | 16 (15-18) | 16 (15-18) | 16 (15-18) | 18 (16-18) | 14 (14-17) | 17 (16-17) | 17 (16-17) | 16 (14-17) | 0.39 |
| **FMS discharge rehabilitation; n (%) <sup>d</sup>** |  |  |  |  |  |  |  |  |  |
| 1 | - | - | - | - | - | - | - | - | 0.70 |
| 2 | 7 (10.9) | 1 (10) | 1 (7.7) | 1 (10) | 1 (12.5) | 2 (22.2) | - | 1 (11.1) |  |
| 3 | 42 (65.6) | 7 (70) | 7 (53.8) | 9 (90) | 5 (62.5) | 4 (44.5) | 4 (80) | 6 (67.7) |  |
| 4 | 14 (21.9) | 2 (20) | 5 (38.5) | - | 1 (12.5) | 3 (33.3) | 1 (20) | 2 (22.2) |  |
| 5 | 1 (1.5) | - | - | - | 1 (12.5) | - | - | - |  |
| **FAC score discharge rehab; n (%)** |  |  |  |  |  |  |  |  |  |
| 0 | - | - | - | - | - | - | - | - | 0.99 |
| 1 | - | - | - | - | - | - | - | - |  |
| 2 | 2 (3) | - | - | - | 1 (11.1) | - | - | 1 (11.1) |  |
| 3 | 2 (3) | - | 1 (7.1) | - | - | - | - | 1 (11.1) |  |
| 4 | 51 (77.3) | 8 (80) | 11 (78.6) | 8 (80) | 7 (77.8) | 7 (77.8) | 4 (80) | 6 (66.7) |  |
| 5 | 11 (16.7) | 2 (20) | 2 (14.3) | 2 (20) | 1 (11.1) | 2 (22.2) | 1 (20) | 1 (11.1) |  |
| BI = Barthel Index, (P)FMS = (Pre-)Fracture Mobility Score, MoCA = Montreal Cognitive Assessment, FAC = Functional Ambulation Categories |  |  |  |  |  |  |  |  |  |
| <sup>a</sup> number of missing = 16, <sup>b</sup> number of missing = 8, <sup>c</sup> number of missing = 3, <sup>d</sup> number of missing = 2 |  |  |  |  |  |  |  |  |  |

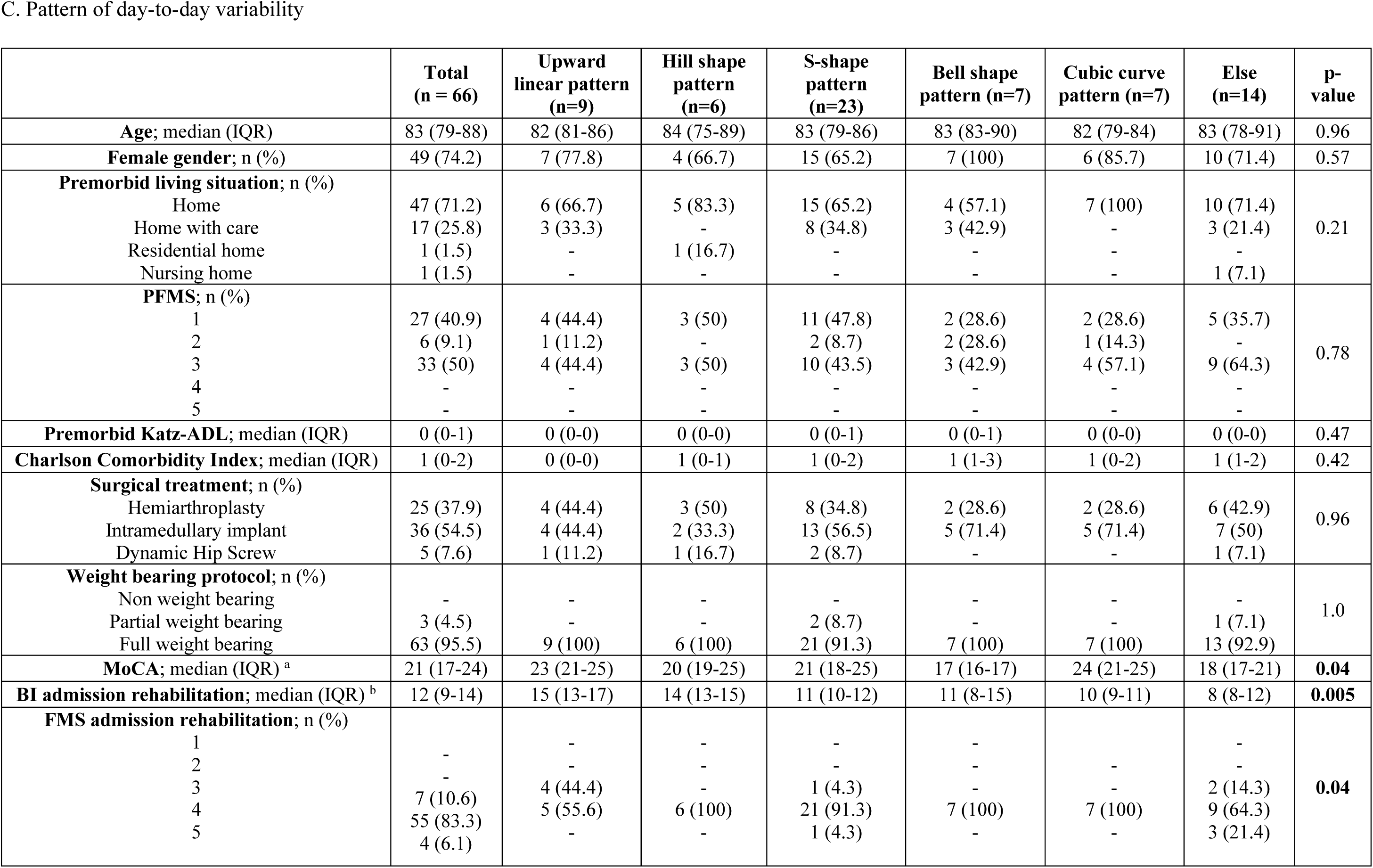
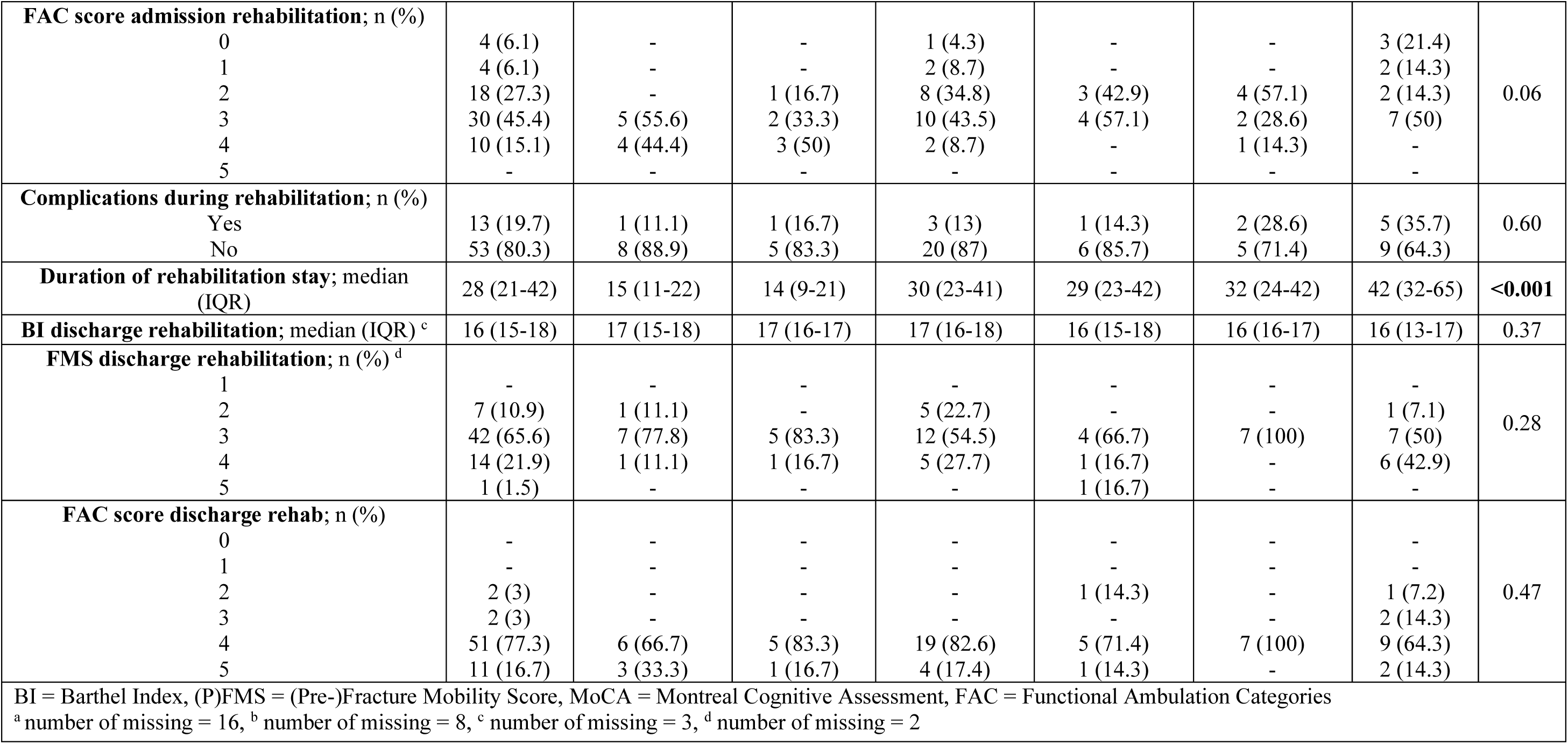

